# Characteristics and predictors of hospitalization and death in the first 9,519 cases with a positive RT-PCR test for SARS-CoV-2 in Denmark: A nationwide cohort

**DOI:** 10.1101/2020.05.24.20111823

**Authors:** Mette Reilev, Kasper Bruun Kristensen, Anton Pottegård, Lars Christian Lund, Jesper Hallas, Martin Thomsen Ernst, Christian Fynbo Christiansen, Henrik Toft Sørensen, Nanna Borup Johansen, Nikolai Constantin Brun, Marianne Voldstedlund, Henrik Støvring, Marianne Kragh Thomsen, Steffen Christensen, Sophie Gubbels, Tyra Grove Krause, Kåre Mølbak, Reimar Wernich Thomsen

## Abstract

**Objective:** To provide population-level knowledge on individuals at high risk of severe and fatal coronavirus disease 2019 (COVID-19) in order to inform targeted protection strategies in the general population and appropriate triage of hospital contacts.

**Design, Setting, and Participants:** Nationwide population-based cohort of all 228.677 consecutive Danish individuals tested (positive or negative) for severe acute respiratory syndrome coronavirus 2 (SARS-CoV-2) RNA from the identification of the first COVID-19 case on February 27^th^, 2020 until April 30^th^, 2020.

**Main Outcomes and Measures:** We examined characteristics and predictors of inpatient hospitalization versus community-management, and death versus survival, adjusted for age-, sex- and number of comorbidities.

**Results:** We identified 9,519 SARS-CoV-2 PCR-positive cases of whom 78% were community-managed, 22% were hospitalized (3.2% at an intensive care unit) and 5.5% had died within 30 days. Median age varied from 45 years (interquartile range (IQR) 31-57) among community-managed cases to 82 years (IQR 75-89) among those who died. Age was a strong predictor of fatal disease (odds ratio (OR) 14 for 70-79-year old, OR 26 for 80-89-year old, and OR 82 for cases older than 90 years, when compared to 50-59-year old and adjusted for sex and number of comorbidities). Similarly, the number of comorbidities was strongly associated with fatal disease (OR 5.2, for cases with ≥4 comorbidities versus no comorbidities), and 82% of fatal cases had at least 2 comorbidities. A wide range of major chronic diseases were associated with hospitalization with ORs ranging from 1.3-1.4 (e.g. stroke, ischemic heart disease) to 2.2-2.7 (e.g. heart failure, hospital-diagnosed kidney disease, chronic liver disease). Similarly, chronic diseases were associated with mortality with ORs ranging from 1.2-1.3 (e.g. ischemic heart disease, hypertension) to 2.4-2.7 (e.g. major psychiatric disorder, organ transplantation). In the absence of comorbidities, mortality was relatively low (5% or less) in persons aged up to 80 years.

**Conclusions and Relevance:** In this first nationwide population-based study, increasing age and number of comorbidities were strongly associated with hospitalization requirement and death in COVID-19. In the absence of comorbidities, the mortality was, however, lowest until the age of 80 years. These results may help in accurate identification, triage and protection of high-risk groups in general populations, i.e. when reopening societies.

## Introduction

Despite worldwide efforts to prevent the spread of the severe acute respiratory syndrome coronavirus 2 (SARS-CoV-2) the derived coronavirus disease 2019 (COVID-19) has become a global pandemic. As per May 18^th^, 2020 COVID-19 has led to more than 4,700,000 confirmed cases and 315,000 deaths worldwide.^1^ In Denmark, the first COVID-19 case was reported on February 27^th^, 2020, and after a few weeks SARS-CoV-2 was widely transmitted in the Danish community.^2^

Hospital-based case series from the early stages of the pandemic have suggested that patient with severe and fatal COVID-19 are likely to be older men with a high burden of comorbid diseases.^3–6 7^ However, a recent analysis from 169 hospitals in 11 countries suggested a median age of only 56 years in fatal cases.^8^ Most previous studies were restricted to hospitals and selected populations in areas where the healthcare systems were overwhelmed by the epidemic. Currently, no studies examined predictors of outcomes in nationwide population-based COVID-19 cohorts in countries with early governmental restrictions and a low burden on the healthcare system. For an unselected nationwide cohort, we describe clinical characteristics and predictors of hospitalization and death for all SARS-CoV-2 PCR-positive cases in Denmark, where early lockdown and surplus healthcare capacity during the epidemic may have influenced the risk of critical disease.

## Methods

In this population-based study of a Danish COVID-19 cohort capturing all individuals with a positive PCR test for SARS-CoV-2 in Denmark, we provide nationwide data on clinical characteristics and predictors of hospitalization and death for all SARS-CoV-2 PCR-positive cases identified from February 27^th^, 2020 to April 30^th^, 2020. For descriptive comparison, we also provide data on clinical characteristics on all individuals with a negative PCR test for SARS-CoV-2 in Denmark. The test-negative subjects can serve as controls in future studies of determinants of contracting COVID-19 infection, using test-negative designs.^9^

### Handling of the epidemic in Denmark

From February 27^th^, 2020 onwards spread of SARS-CoV-2 in Denmark was observed within clusters and mainly suspected symptomatic COVID-19 cases with a relevant travel history (mainly from China and Italy) were tested. As per March 12^th^ community transmission was observed and it was decided to shift from a containment to a mitigation strategy, where testing of patients who had suspected COVID-19 requiring hospital admission was prioritized and contact tracing with quarantine was stopped. The government instituted a comprehensive lockdown of the country on March 13^th^. On 18^th^ March testing of frontline health care workers in critical functions who had respiratory symptoms was possible and from late March onwards, test capacity was gradually upscaled to include testing of individuals with mild to moderate respiratory symptoms suspicious of COVID-19, as well as broader screening of healthcare professionals. A controlled and gradual reopening of selected sectors of the country was initiated on April 15^th^.

### The Danish SARS-CoV-2 cohort

We established the study cohort using data on SARS-CoV-2 PCR results from the Danish Microbiology Database.^10, 11^ Using the unique personal identifier assigned to all Danish citizens, the study cohort was linked to the Danish administrative and health registries.^12–17^ We obtained complete information on use of prescription drugs, history of hospitalizations and comorbidities, authorized health care worker status, admission to ICU, and date of death if any (for definitions of variables, see **Supplementary Table 1**).

A case was defined as an individual tested one or more times with at least one positive PCR test result for SARS-CoV-2 performed on oro- and nasopharyngeal swaps and/or on respiratory tract secretions and aspirates. The date of the first positive PCR test was used as the index date while individuals with negative SARS-CoV-2 PCR tests were included by the date of their first negative test. Hospital admissions due to COVID-19 were defined as continuous in-hospital stays with a duration of 12 hours or longer occurring up to 14 days after the index date. ICU treatment was defined as intensive care treatment from two days before the index date to 14 days after and was identified using procedure codes in the Danish National Patient Registry^12^ or by direct reporting from the Danish Regions to Statens Serum Institut. Mortality was defined as deaths occurring from two days before the index date to 30 days after and validated in the Danish Cause of Death Registry.^13^ Real time data updates were available on the entire cohort. We included PCR positive cases with an index date prior to April 30^th^, 2020 in the main analysis. Since follow-up data availability ended May 15 and a minor proportion of PCR-positive cases thus had less than 30 days of follow-up (i.e., cases diagnosed between April 16 and April 30), we performed a sensitivity analysis in which we assessed predictors of 30-day mortality only including individuals with an index date prior to April 15^th^, 2020.

### Analysis

We first assessed the number of SARS-CoV-2 PCR positive cases in Denmark as well as the number of individuals tested PCR negative for SARS-CoV-2. Second, we described clinical characteristics for individuals with negative SARS-CoV-2 PCR results and for PCR positive cases further stratified by disease severity, i.e. cases who were managed in the community, cases requiring hospitalization, cases requiring ICU admission, and cases who died within 30 days (inside or outside hospitals). Third, we charted the proportion of hospitalized cases and cases who died within 30 days, specified by age, and assessed predictors of hospitalization and death by estimating crude, and age- and sex-adjusted odds ratios (ORs) using logistic regression for the associations between single comorbidities and hospitalization and death within 30 days. We chose a logistic regression over a conventional Cox regression as we observed a high number of patients who died very shortly after being tested positive. A survival analysis, like a Cox regression, would put undue emphasis on the time interval between the positive test and death, whereas a logistic regression would merely reflect predictors of whether the patient died or not. As a measure of relative risk, the OR is an overestimate when the risk of an outcome is common. To examine if the outcome associations with age and sex depended on the related burden of comorbidity, we additionally adjusted age and sex ORs for number of comorbidities. Comorbidities were defined as an ever-recording of chronic lung disease, hypertension, ischemic heart disease, heart failure, atrial fibrillation, stroke, diabetes, dementia, any cancer, chronic liver disease, hospital-diagnosed kidney disease, alcohol abuse, substance abuse, major psychiatric disorders, organ transplantation, overweight and obesity, and/or rheumatoid arthritis/connective tissue disease whereas the number of comorbidities was defined as the total number of any of these co-existing conditions. In a supplementary analysis, ORs were estimated for single comorbidities while adjusting for age, sex, and additionally for total number of comorbidities. Finally, we investigated clinical characteristics of patients diagnosed during different phases of the epidemic in Denmark, defined as the containment phase, the mitigation phase, and during the gradual reopening.

### Other

According to Danish law, studies based entirely on registry data do not require approval from an ethics review board.^17^ Due to legal reasons, individual level raw data from Danish administrative and health registries cannot be shared by the authors.

## Results

We identified 9,519 cases with SARS-CoV-2 detected by PCR and 219,158 individuals with a negative PCR test in the Danish SARS-CoV-2 cohort from February 27^th^, 2020 to April 30^th^, 2020. The number of new SARS-CoV-2 PCR positive cases peaked at around 470 cases per day 5-6 weeks after the identification of the first case. The number of new positive cases, however, correlated closely with the number of individuals being tested, thus mainly reflecting changes in instituted test strategies (**Figure 1**).

**Figure 1.**
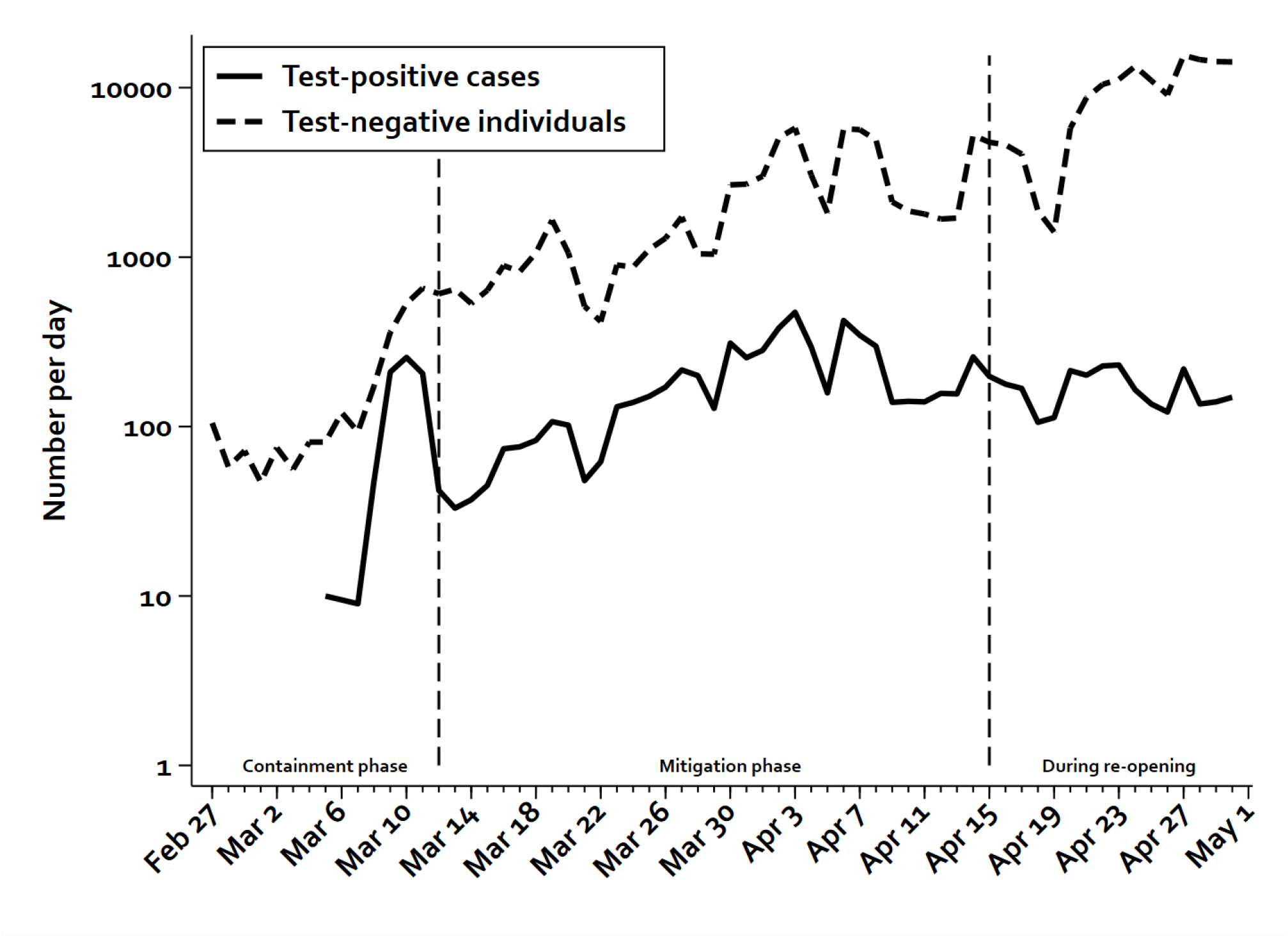
New SARS-CoV-2 PCR positive cases and the number of individuals tested negative for SARS-CoV-2 per day during the stages of the ongoing epidemic. The dotted lines illustrate the shift from containment to mitigation strategy as well as to the reopening of the society. Note logarithmic y-axis.

In general, we only observed minor differences in age, sex, medical history and prior drug use between PCR positive cases and test-negative individuals (**Table 1**). Among all SARS-CoV-2 PCR positive cases in Denmark, 22% required hospitalization while 3.2% were admitted to an ICU and 5.5% had a fatal course of disease within 30 days from the positive test (**Table 1**). Of those who died, 22% were managed in the community i.e., by definition they did not have an in-hospital admission longer than 12 hours within 14 days after the index date (**Supplementary Table 2**). The majority of hospitalized cases were admitted on the date of the positive PCR test (59%) (**Supplementary Figure 1**). Forty-two percent of all PCR positive cases were men, increasing to 74% among cases admitted to an ICU and 57% among cases who died within 30 days of the positive test (**Table 1**). When adjusted for age and number of comorbidities, ORs for hospitalization and death were 1.8 and 2.1, respectively for men (**Table 2**).

**Table 1.**
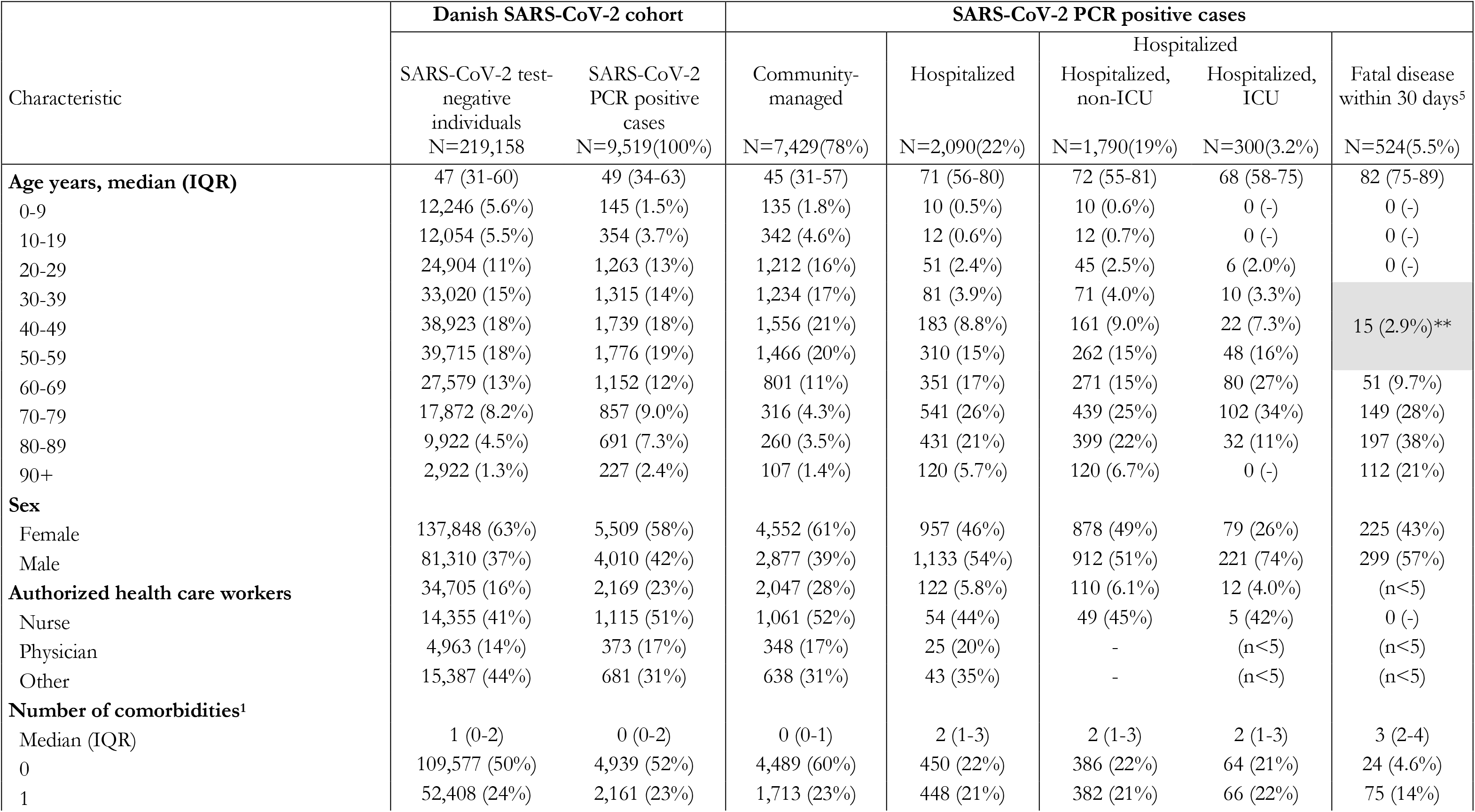
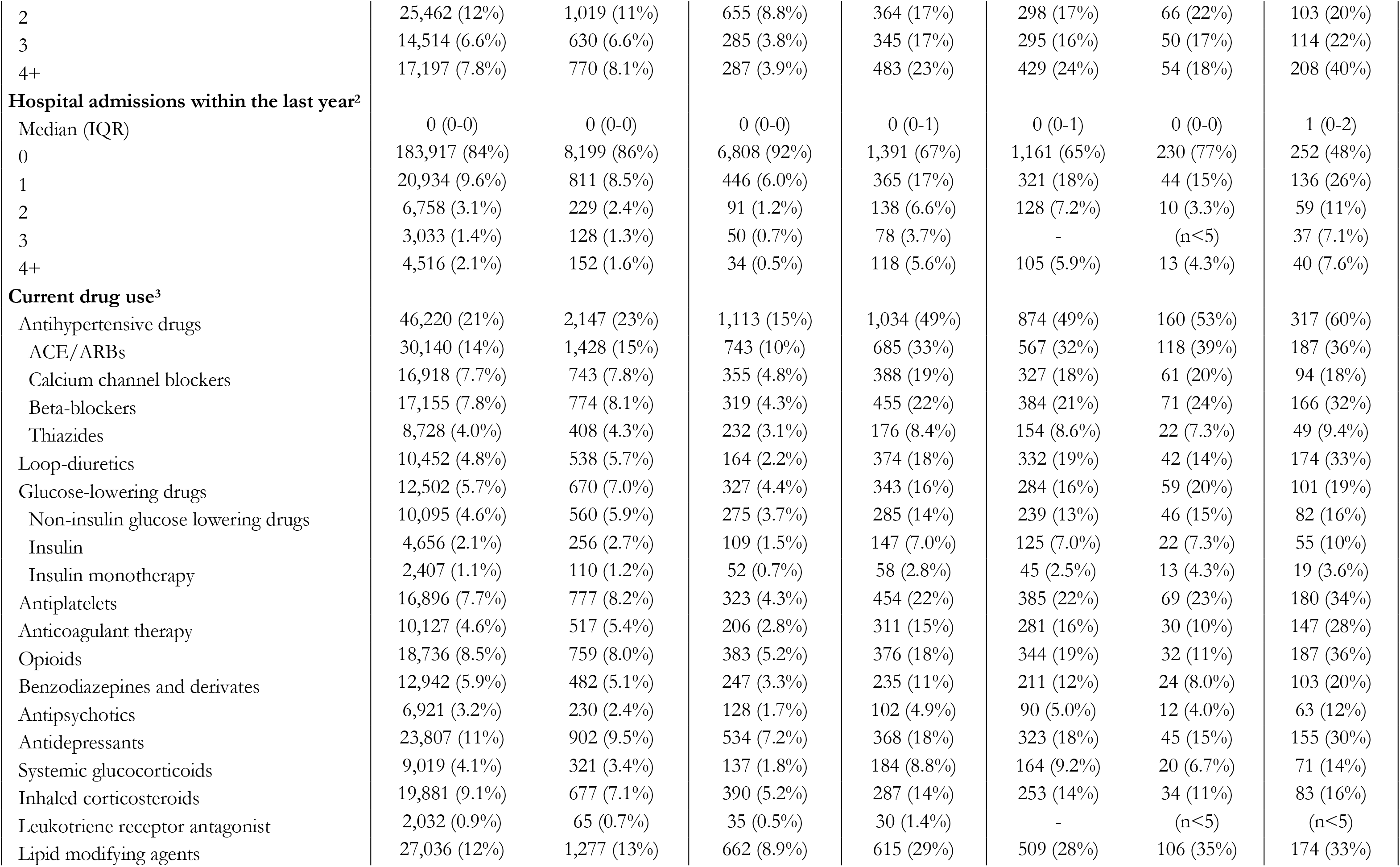
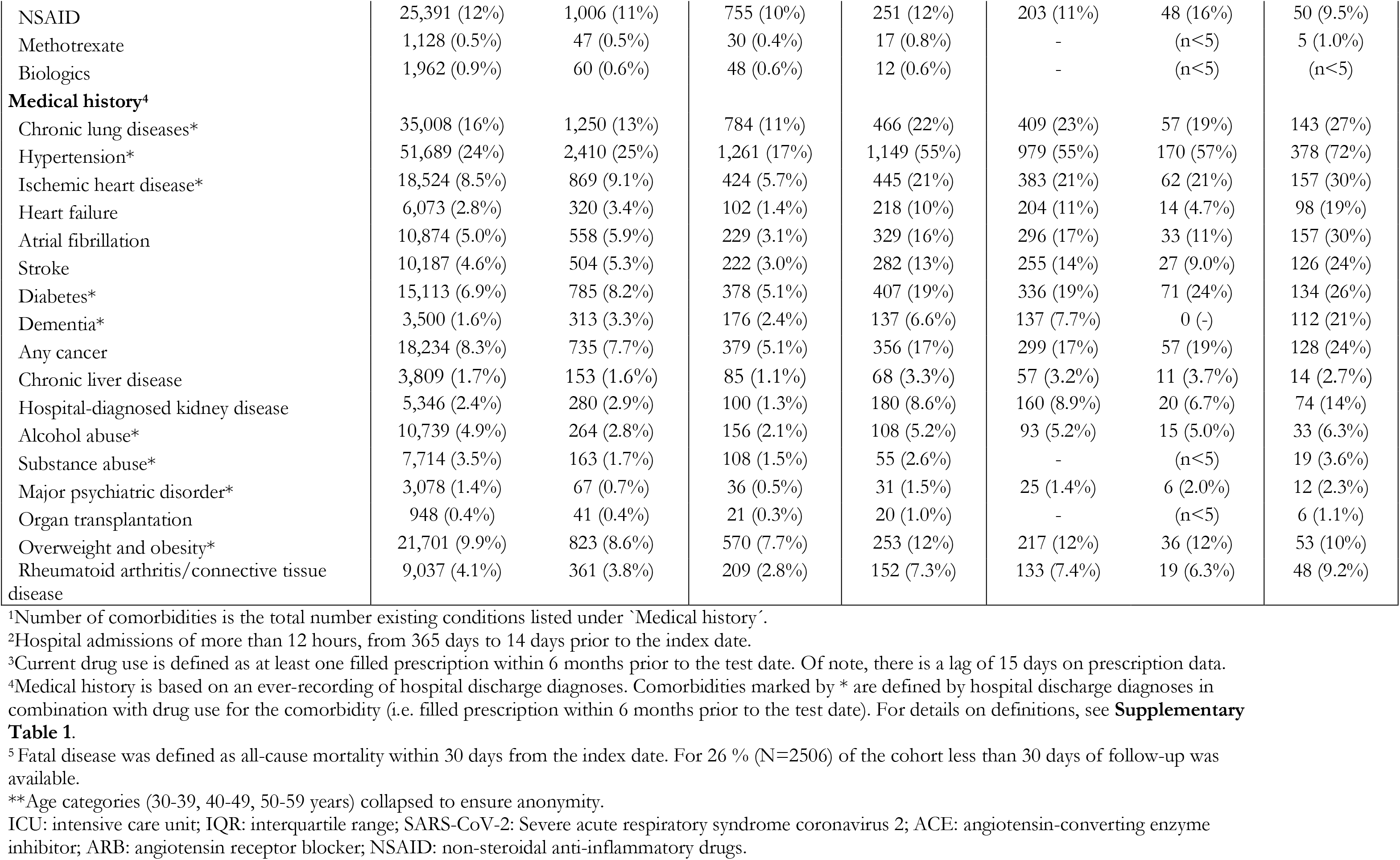
Baseline characteristics for the overall Danish SARS-CoV-2 cohort and specified by whether the infection was community-managed or led to any hospitalization, hospitalization without ICU-admission, hospitalization with ICU-admission, or death.

**Table 2.** Predictors of hospitalization and having a fatal course among SARS-CoV-2 PCR positive cases.

| Characteristics | Hospitalization |  |  | Death within 30 days<sup>5</sup> |  |  |
| --- | --- | --- | --- | --- | --- | --- |
|  | Crude OR (95%CI) | Age- and sex-adjusted OR (95%CI) | Age-, sex-, and number of comorbidities adjusted OR (95%CI)<sup>1</sup> | Crude OR (95%CI) | Age- and sex-adjusted OR (95%CI) | Age-, sex-, and number of comorbidities adjusted OR (95%CI)<sup>1</sup> |
| **Age, years<sup>1</sup>** |  |  |  |  |  |  |
| 0-9 | 0.4 (0.2-0.7) | 0.3 (0.2-0.6) | 0.4 (0.2-0.9) | NA | NA | NA |
| 10-19 | 0.2 (0.1-0.3) | 0.2 (0.1-0.3) | 0.2 (0.1-0.4) | NA | NA | NA |
| 20-29 | 0.2 (0.1-0.3) | 0.2 (0.1-0.3) | 0.3 (0.2-0.3) | NA | NA | NA |
| 30-39 | 0.3 (0.2-0.4) | 0.3 (0.2-0.4) | 0.4 (0.3-0.5) | NA | NA | NA |
| 40-49 | 0.6 (0.5-0.7) | 0.6 (0.5-0.7) | 0.6 (0.5-0.8) | NA | NA | NA |
| 50-59 | 1.00 (ref.) | 1.00 (ref.) | 1.00 (ref.) | 1.00 (ref.) | 1.00 (ref.) | 1.00 (ref.) |
| 60-69 | 2.1 (1.7-2.5) | 2.0 (1.7-2.4) | 1.6 (1.4-2.0) | 5.8 (3.2-10.6) | 5.4 (3.0-9.9) | 4.2 (2.3-7.6) |
| 70-79 | 8.1 (6.7-9.7) | 7.7 (6.3-9.2) | 4.8 (3.9-5.9) | 26.5 (15.2-46.1) | 24.0 (13.8-41.9) | 13.5 (7.6-24.0) |
| 80-89 | 7.8 (6.4-9.5) | 8.0 (6.6-9.8) | 4.5 (3.6-5.6) | 50.2 (28.9-87.1) | 51.0 (29.4-88.7) | 26.4 (14.9-46.8) |
| 90+ | 5.3 (4.0-7.1) | 5.9 (4.4-7.9) | 3.5 (2.6-4.7) | 122.6 (68.2-220.4) | 146.6 (81.0-265.1) | 82.2 (44.7-151.1) |
| **Sex<sup>1</sup>** |  |  |  |  |  |  |
| Female | 1.00 (ref.) | 1.00 (ref.) | 1.00 (ref.) | 1.00 (ref.) | 1.00 (ref.) | 1.00 (ref.) |
| Male | 1.9 (1.7-2.1) | 1.8 (1.6-2.0) | 1.8 (1.6-2.0) | 1.9 (1.6-2.3) | 2.1 (1.7-2.6) | 2.1 (1.7-2.7) |
| **Authorized health care workers** |  |  |  |  |  |  |
| Non-health care worker | 1.00 (ref.) | 1.00 (ref.) | 1.00 (ref.) | 1.00 (ref.) | 1.00 (ref.) | 1.00 (ref.) |
| Nurse | 0.1 (0.1-0.2) | 0.4 (0.3-0.5) | 0.4 (0.3-0.6) | NA | NA | NA |
| Physician | 0.2 (0.1-0.3) | 0.4 (0.3-0.6) | 0.4 (0.3-0.7) | NA | NA | NA |
| Other | 0.2 (0.1-0.3) | 0.4 (0.3-0.6) | 0.4 (0.3-0.6) | NA | NA | NA |
| **Number of comorbidities<sup>2</sup>** |  |  |  |  |  |  |
| 0 | 1.00 (ref.) | 1.00 (ref.) |  | 1.00 (ref.) | 1.00 (ref.) |  |
| 1 | 2.6 (2.3-3.0) | 1.7 (1.4-2.0) |  | 7.4 (4.6-11.7) | 2.4 (1.5-4.0) |  |
| 2 | 5.5 (4.7-6.5) | 2.0 (1.7-2.5) |  | 23.0 (14.7-36.1) | 2.9 (1.8-4.8) |  |
| 3 | 12.1 (10.0-14.5) | 3.2 (2.6-3.9) |  | 45.2 (28.9-70.9) | 3.8 (2.3-6.3) |  |
| 4+ | 16.8 (14.1-20.0) | 3.6 (2.9-4.5) |  | 75.8 (49.2-116.7) | 5.2 (3.2-8.3) |  |
| **Hospital contacts within the last year<sup>3</sup>** |  |  |  |  |  |  |
| 0 | 1.00 (ref.) | 1.00 (ref.) | 1.00 (ref.) | 1.00 (ref.) | 1.00 (ref.) | 1.00 (ref.) |
| 1 | 4.0 (3.4-4.7) | 2.2 (1.8-2.6) | 1.9 (1.6-2.3) | 6.4 (5.1-7.9) | 2.1 (1.7-2.8) | 1.8 (1.4-2.4) |
| 2 | 7.4 (5.7-9.7) | 3.3 (2.4-4.5) | 2.7 (2.0-3.8) | 10.9 (7.9-15.1) | 2.9 (2.0-4.2) | 2.5 (1.7-3.6) |
| 3 | 7.6 (5.3-10.9) | 2.6 (1.8-4.0) | 2.1 (1.4-3.1) | 12.8 (8.6-19.2) | 2.8 (1.8-4.4) | 2.2 (1.4-3.5) |
| 4+ | 17.0 (11.5-25.0) | 7.7 (5.0-11.9) | 5.4 (3.5-8.4) | 11.3 (7.7-16.5) | 2.9 (1.9-4.5) | 2.2 (1.4-3.4) |
| **Medical history<sup>4</sup>** |  |  |  |  |  |  |
| Chronic lung diseases* | 2.4 (2.1-2.8) | 1.7 (1.5-2.0) |  | 2.7 (2.2-3.3) | 1.4 (1.1-1.8) |  |
| Hypertension* | 6.0 (5.4-6.6) | 1.8 (1.5-2.0) |  | 8.9 (7.3-10.8) | 1.3 (1.1-1.7) |  |
| Ischemic heart disease* | 4.5 (3.9-5.2) | 1.4 (1.2-1.7) |  | 5.0 (4.1-6.1) | 1.2 (0.9-1.5) |  |
| Heart failure | 8.4 (6.6-10.6) | 2.2 (1.7-2.9) |  | 9.1 (7.0-11.8) | 1.7 (1.3-2.2) |  |
| Loop-diuretic use** | 9.7 (8.0-11.7) | 2.4 (2.0-3.0) |  | 11.8 (9.6-14.5) | 2.0 (1.6-2.5) |  |
| Atrial fibrillation | 5.9 (4.9-7.0) | 1.4 (1.1-1.7) |  | 9.2 (7.4-11.3) | 1.6 (1.2-2.0) |  |
| Stroke | 5.1 (4.2-6.1) | 1.3 (1.0-1.6) |  | 7.2 (5.8-9.0) | 1.4 (1.1-1.8) |  |
| Diabetes* | 4.5 (3.9-5.2) | 1.9 (1.6-2.2) |  | 4.4 (3.6-5.4) | 1.6 (1.3-2.1) |  |
| Non-insulin glucose lowering drug use** | 4.1 (3.5-4.9) | 1.8 (1.4-2.2) |  | 3.3 (2.6-4.3) | 1.3 (1.0-1.8) |  |
| Any insulin use** | 5.1 (3.9-6.5) | 2.1 (1.6-2.8) |  | 5.1 (3.8-7.0) | 2.0 (1.4-2.9) |  |
| Insulin monotherapy use** | 4.0 (2.8-5.9) | 2.2 (1.4-3.4) |  | 3.7 (2.2-6.1) | 1.6 (0.9-2.8) |  |
| Dementia* | 2.9 (2.3-3.6) | 0.5 (0.4-0.7) |  | 11.9 (9.3-15.3) | 1.9 (1.4-2.5) |  |
| Any cancer | 3.8 (3.3-4.5) | 1.3 (1.1-1.6) |  | 4.5 (3.6-5.5) | 1.3 (1.0-1.7) |  |
| Chronic liver disease | 2.9 (2.1-4.0) | 2.5 (1.7-3.6) |  | 1.7 (1.0-3.1) | 1.6 (0.8-3.0) |  |
| Hospital-diagnosed kidney disease | 6.9 (5.4-8.9) | 2.7 (2.0-3.6) |  | 7.0 (5.3-9.3) | 2.0 (1.4-2.7) |  |
| Alcohol abuse* | 2.5 (2.0-3.3) | 1.7 (1.2-2.3) |  | 2.5 (1.8-3.7) | 1.6 (1.1-2.5) |  |
| Substance abuse* | 1.8 (1.3-2.5) | 1.4 (0.9-2.0) |  | 2.3 (1.4-3.8) | 1.8 (1.0-3.1) |  |
| Major psychiatric disorder* | 3.1 (1.9-5.0) | 2.1 (1.2-3.8) |  | 3.8 (2.0-7.2) | 2.4 (1.1-5.0) |  |
| Benzodiazepines and derivates use** | 3.7 (3.1-4.4) | 1.7 (1.3-2.1) |  | 5.6 (4.4-7.1) | 2.0 (1.5-2.6) |  |
| Antipsychotic use** | 2.9 (2.2-3.8) | 1.4 (1.0-1.8) |  | 7.2 (5.3-9.8) | 3.6 (2.5-5.2) |  |
| Antidepressant use** | 2.8 (2.4-3.2) | 1.3 (1.1-1.5) |  | 4.6 (3.8-5.7) | 1.7 (1.4-2.2) |  |
| Organ transplantation | 3.4 (1.8-6.3) | 2.7 (1.3-5.4) |  | 3.0 (1.2-7.1) | 2.7 (1.0-7.5) |  |
| Overweight and obesity* | 1.7 (1.4-1.9) | 2.0 (1.7-2.4) |  | 1.2 (0.9-1.6) | 1.5 (1.1-2.1) |  |
| Rheumatoid arthritis/connective tissue disease | 2.7 (2.2-3.4) | 1.4 (1.1-1.8) |  | 2.8 (2.0-3.8) | 1.0 (0.7-1.5) |  |

<sup>1</sup>Age was adjusted for sex and number of comorbidities while sex was adjusted for age and number of comorbidities.
<sup>2</sup>Number of comorbidities is the total number of coexisting conditions listed under 'Medical history'.
<sup>3</sup>Hospital admissions of more than 12 hours, from 365 days to 14 days prior to the index date.
<sup>4</sup>Medical history is based on an ever-recording of hospital discharge diagnoses. Comorbidities marked by \* are defined by hospital discharge diagnoses in combination with drug use for the comorbidity (i.e. filled prescription within 6 months prior to the test date). \*\* denotes exclusive use of drugs that are close markers of specific underlying comorbidities, assessed independently of presence or absence of hospital diagnoses for the comorbidity. For details on definitions, see **Supplementary Table 1**.
<sup>5</sup>Death was defined as all-cause mortality within 30 days from the index date.
OR: odds ratio; SARS-CoV-2: Severe acute respiratory syndrome coronavirus 2.

Among all PCR positive cases, the median age was 49 years (IQR 34-63), varying from 45 years (IQR 31-57) among cases who did not require hospitalization to 82 years (IQR 75-89) among those who died (**Table 1**). The proportion of SARS-CoV-2 PCR positive cases requiring hospitalization increased substantially with age to more than 60% among cases older than 70 years (**Figure 2**). Similarly, the proportion of PCR positive cases with a fatal course increased from 17% by the age of 70-79 years to 29% by the age of 80-89 years (**Figure 2**). When adjusting for sex and number of comorbidities, increasing age was a very strong predictor of fatal disease (OR 14 for 70-79-year old, OR 26 for 80-89-year old, and OR 82 for cases older than 90 years, when compared to middle-aged adults, 50-59 years (**Table 2**).

**Figure 2.**
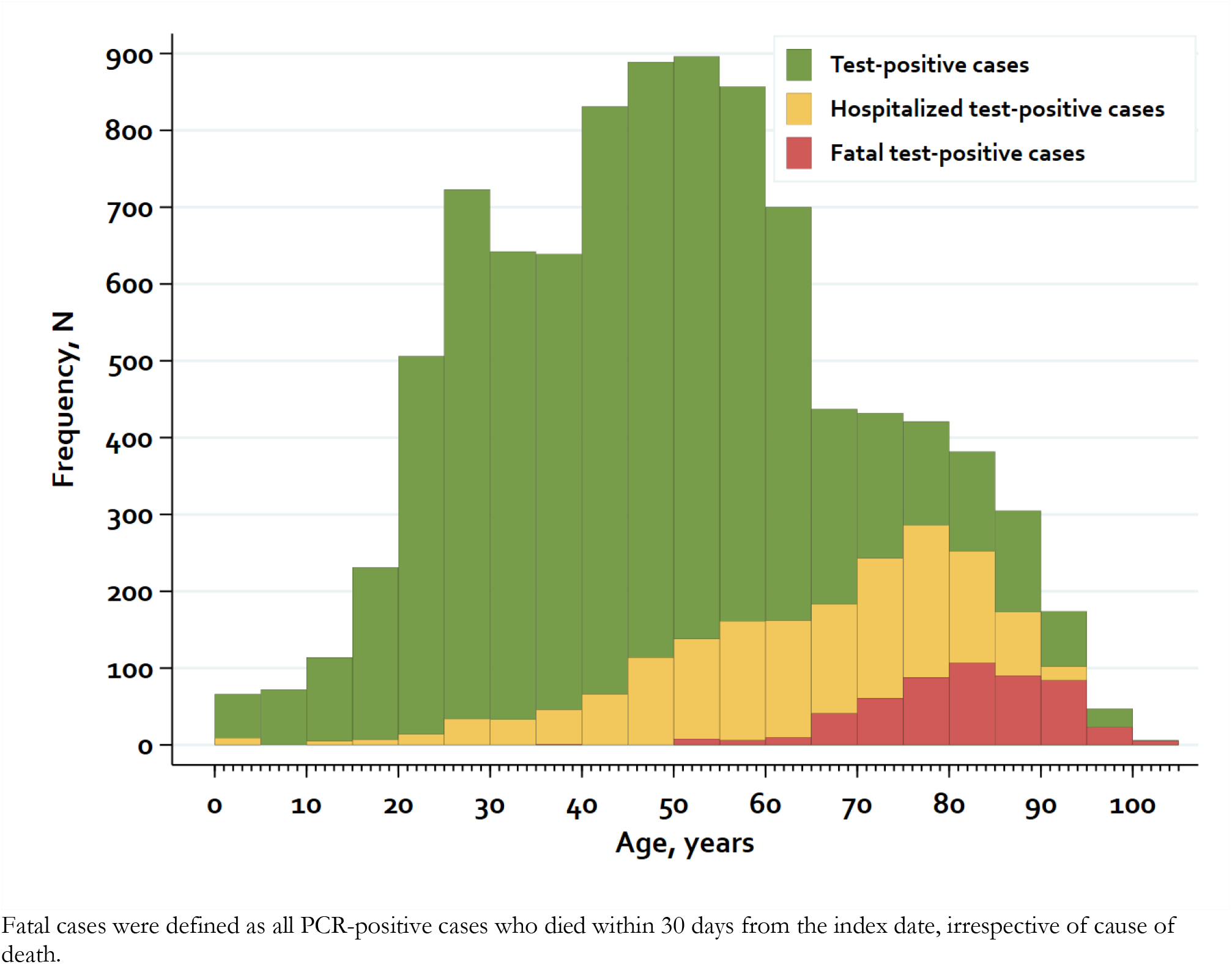
Distribution of hospitalization and death according to age in all SARS-CoV-2 PCR positive cases.

In general, comorbidities were more frequent among PCR positive hospitalized and fatal cases. Thus, 17% of community-managed cases had 2 or more comorbidities, while the corresponding proportion was 57% among hospitalized and 82% among fatal cases. Similarly, the proportion of individuals who had been hospitalized at least once during the last year was higher among hospitalized cases (33%) and fatal cases (52%) than among PCR positive cases managed in the community (8%) (**Table 1**).

The most frequent comorbidities among hospitalized cases were hypertension (55%), COPD (22%), ischemic heart disease (21%), and diabetes (19%) (**Table 1**). After adjustment for higher age and sex, the association of many comorbidities with hospitalization risk reduced considerably (e.g. for dementia, from OR 2.9 to OR 0.5). However, the other comorbidities remained predictive of COVID-19 hospitalization, ranging from OR 1.3 for e.g. cancer or stroke, OR 1.4 for atrial fibrillation or ischemic heart disease, to OR 1.7 for chronic lung disease, OR 1.8 for hypertension, OR 1.9 for diabetes, and peaking at OR 2.7 for hospital-diagnosed kidney disease and organ transplantation (**Table 2**). In large, this pattern was also evident among fatal cases, though the absolute prevalence of comorbidities was higher in fatal than hospitalized cases (**Table 1 and 2**). Among PCR positive cases with 4 or more comorbidities, the ORs for hospitalization was 3.6 and 5.2 for death compared to PCR positive cases without any comorbidities (**Table 2**).

Mortality increased substantially when combining increasing age with increasing number of comorbidities (**Figure 3**). Among 60-69-year old cases and 70-79-year old cases with no comorbidities the mortality was relatively low (1% and 5%), however, increasing to 9% and 28%, respectively among those with ≥4 comorbidities. Among the oldest old cases mortality was high regardless of number of comorbidities (**Figure 3**).

**Figure 3.**
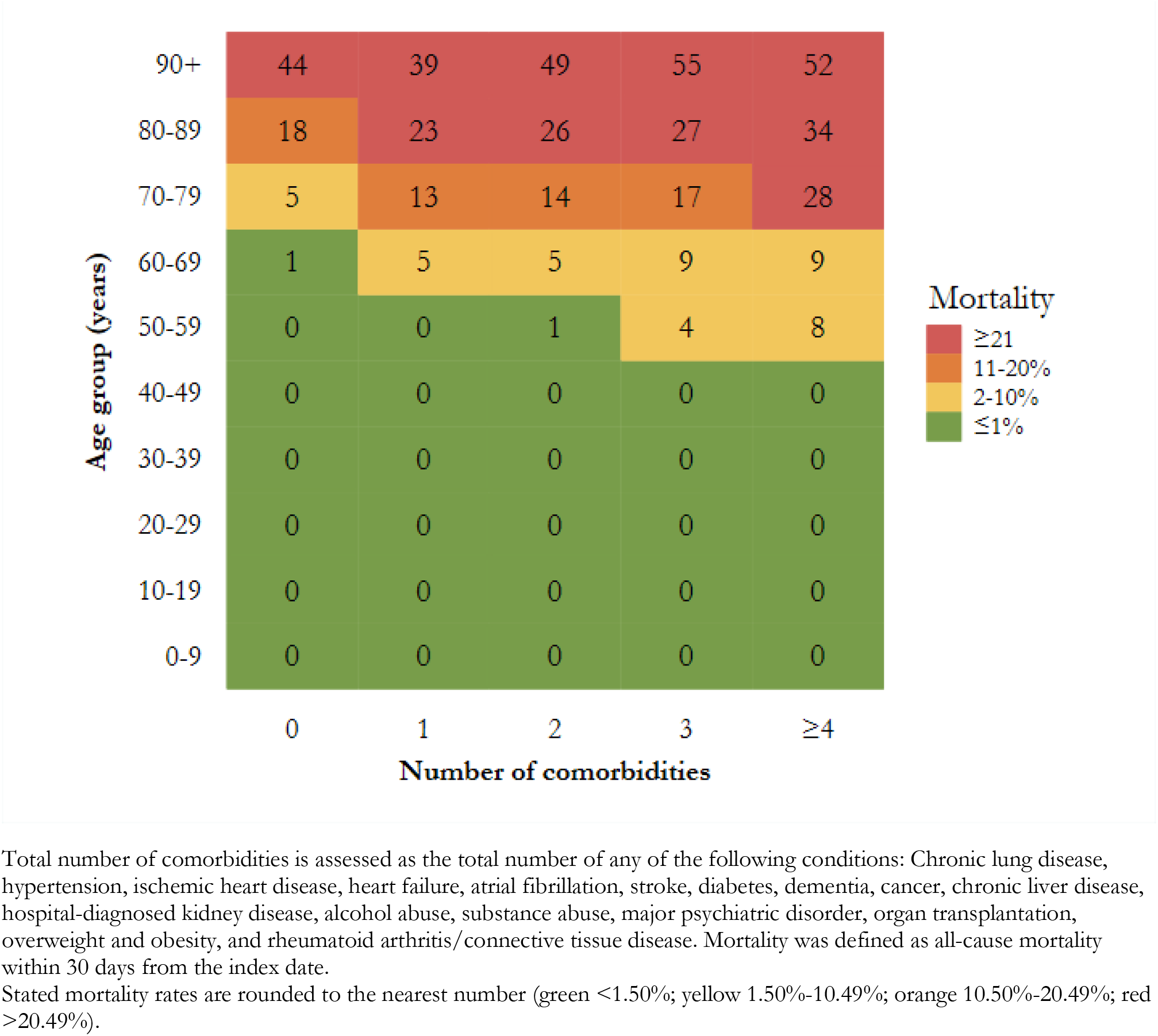
Heatmap illustrating mortality among SARS-CoV-2 PCR positive cases, specified by age and number of comorbidities.

When we only included individuals with minimum follow-up of 30 days (index date prior to April 15th, 2020), the ORs for different predictors did not differ materially (data not shown).

When further adjusting for total number of comorbidities, the ORs for patients with individual comorbidities declined considerably (**Supplementary Table 3**), suggesting that multimorbidity and frailty in patients with e.g. hypertension, diabetes, or cardiopulmonary disease may be a key driver of the observed associations.

Authorized healthcare workers comprised 23% of all PCR positive case (**Table 1**). Of these, 122 cases (5.6%) required hospital admission, 12 cases (0.6%) required ICU admission, and <5 cases (<0.2%) died within 30 days of the positive test.

Patient characteristics of all PCR positive cases changed markedly during the different stages of the epidemic. Thus, the proportion of women increased from 32% in the initial stage (when travelers were frequently tested) to 62% during the reopening stage (when healthcare workers were frequently tested).

The median age was highest during the mitigation phase (52 years, IQR 38-66) where predominantly individuals requiring hospitalization were tested (**Supplementary Table 4**).

## Discussion

In this nationwide cohort of SARS-CoV-2 PCR positive cases and test-negative individuals from the general population in Denmark, we found that older age (e.g., >70 years), male sex, and number of comorbidities were risk factors for hospitalization and death. In the absence of comorbidities, the mortality was, however, 5% or below until the age of 80 years. After controlling for age and sex, virtually all comorbidities that were prevalent in our population, including e.g. hypertension, heart or lung disease, obesity, and diabetes were associated with severe disease or death from COVID-19. Particularly strong associations were observed for hospital-diagnosed kidney disease, severe psychiatric disorder, and organ transplantation.

To our knowledge, this is the first nationwide study in which clinical characteristics and predictors of hospitalization and death of SARS-CoV-2 PCR positive cases are investigated at the population level in a country controlling the outbreak with early restriction leading to surplus health care capacity during the epidemic. The register-based approach and Denmark’s universal health care system is a major strength of this study, since the Danish administrative and health registries allow complete nationwide capture of an unselected cohort of all individuals tested for SARS-CoV-2 without restricting to those treated at hospitals and irrespective of socio-economic differences. Population-based registries allowed for complete, independent individual-level ascertainment of all previous hospital contacts and prescription drug use, overcoming limitations of missing data in previous reports. Such complete mapping of medical history allowed us to establish any effect modification caused by increasing age and number of comorbidities.

The existence of associations between largely any comorbidities and the risk of hospitalization and death due to COVID-19 in our study is in accordance with previous studies of both COVID-19-patients^18,19^ and patients with severe influenza,^20^ thus suggesting resemblances between COVID-19 and other severe respiratory infections with regards to populations at risk. Among the most frequent comorbidities in our population, hypertension, obesity and diabetes seemed to be clear predictors of both hospitalization and fatal disease, which corroborates previous hospital-based outcome studies ^21,22^ and underscores the probable importance of metabolic health in COVID-19 outcomes.^23^ Our data add important new knowledge on the possible role of hospital-diagnosed kidney disease and organ transplantation as strong risk factors, and furthermore suggest that people with alcohol/substance abuse and psychiatric illness may be an especially vulnerable group, possibly in line with the socioeconomic disparities that have been observed during the COVID-19 epidemic.^24,25^

Importantly, the assessment of predictors in our study was performed without having any prespecified hypotheses. We did not aim at estimating causal effects, but rather to identify factors that could help us identify people at high risk of a severe COVID-19 course. The potential causal associations of specific individual diseases with COVID-19 outcomes, including the potentially strong association observed for e.g. metabolic diseases should be analyzed in future epidemiological studies designed to evaluate causal effects, including detailed, hypothesis-specific confounder assessment. Any associations observed in this study should therefore be interpreted with caution, and not as evidence of causality. Moreover, the threshold for diagnosing comorbidities as well as COVID-19 may differ across age groups. Among elderly multimorbid persons, Berksonian-like bias may have caused an overestimation of COVID-19-outcome associations, if some hospital admissions were primarily related to worsening underlying comorbidities rather than infection, and then led to testing and coincident detection of SARS-CoV-2.

It is of note that the different test strategies instituted in Denmark during the epidemic are crucial for the observed characteristics of individuals with confirmed SARS-CoV-2 infection. In the early stages of the pandemic, the national test strategy in Denmark was directed at those who were most sick and potentially in need of medical care, which may have contributed to the rather high proportion of cases hospitalized in our study. Also, the high absolute number of PCR positive healthcare professionals may reflect widespread testing in this group to track down and limit in-hospital contamination, rather than a particularly high level of contamination of healthcare professionals. Furthermore, the source of transmission is unknown. It is a limitation in our study that the subgroup of healthcare professionals only included individuals authorized as healthcare personnel, i.e. mainly nurses and doctors but not professions where an authorization is not required e.g. hospital porters and nurse assistants. Further, data on whether the healthcare professionals were involved in clinical work or working in other settings was not available, thus some health care professionals without current patient contact were included.

In accordance with previous descriptive studies, older men with comorbidities dominated the subgroup of cases with severe or fatal disease. ^26,27^ Additionally, we found that men had a 2.1-fold risk of death even when adjusting for age and number of comorbidities, thus suggesting that the higher risk of severe or fatal disease among men cannot be explained by a higher burden of comorbidities in its entirety. In our study, cases admitted to an ICU were markedly younger and less comorbid than cases with a fatal course, which is likely explained by a qualified triage of patients suitable for intensive care treatment.

Also, COVID-19 cases with a fatal course who were not hospitalized (22% of fatalities) were older and more likely to have dementia than those who died following hospitalization, thus similarly suggesting triage of cases according to age and frailty. Of note, the median age of 82 years at death from COVID-19 in our study is almost identical to the median age at death (81 years) of general population members in Denmark.^28^

## Conclusion

In this first nationwide population-based study, increasing age, sex, and number and type of comorbidities were closely associated with hospitalization requirement and death in SARS-CoV-2 PCR positive cases. In the absence of comorbidities, the mortality was, however, lowest until the age of 80 years. These results may help in accurate identification, triage and protection of high-risk groups in general populations, i.e. when reopening societies.

## Data Availability

Due to legal reasons, individual level raw data from the Danish Administrative and Health Registry cannot be shared by the authors.

## Acknowledgements

We would like to thank all the Departments of Clinical Microbiology throughout Denmark and the data integration og analyse (DIAS), Infektionsberedskabet, and Jonas Kähler and Karsten Dalsgaard Bjerre from Statens Serum Institut, Copenhagen for their contribution of data in this study.

## Contributions

All authors designed the study, interpreted the data, revised the manuscript, and approved the final version of the manuscript. Martin Thomsen Ernst cleaned and analyzed the data. Kasper Bruun Kristensen validated the code used for data cleaning and analysis.

## Disclosure statements

KBK, NBJ, MAV, SC, NCB, CFC, JH, MTE, KM, TGK, SG and MKT declare no conflicts of interest. RWT, and HTS declare no personal conflicts of interest. The Department of Clinical Epidemiology is involved in studies with funding from various companies as research grants to and administered by Aarhus University. None of these studies are related to the current study.

HS reports personal fees from Bristol-Myers Squibb, personal fees from Novartis, personal fees from Roche, outside the submitted work.

AP report participation in research funded by Alcon, Almirall, Astellas, AstraZeneca, Boehringer-Ingelheim, Novo Nordisk, Servier and LEO Pharma, all with funds paid to the institution where they were employed (no personal fees) and with no relation to the work reported in this paper.

LCL reports participation in research projects funded by Menarini Pharmaceutical and LEO Pharma, with funds paid to the institution where he was employed (no personal fees) and with no relation to the work reported in this paper.

MR reports participation in research projects funded by LEO Pharma, with funds paid to the institution where she was employed (no personal fees) and with no relation to the work reported in this paper.

## Funding

None

## Notes

### Author Declarations

According to Danish law, studies based entirely on registry data do not require approval from an ethics review board.

